# Optimized decision support for selection of transoral robotic surgery or (chemo)radiation: Quantified pre-therapy risk stratification for patient-reported and clinician-graded swallowing impairment and toxicity

**DOI:** 10.1101/2021.06.12.21258794

**Authors:** Mehdi Hemmati, Carly Barbon, Abdallah S.R. Mohamed, Lisanne V. van Dijk, Amy C. Moreno, Neil D. Gross, Ryan P. Goepfort, Stephen Y. Lai, Katherine A. Hutcheson, Andrew Schaefer, Clifton D. Fuller, for the MD Anderson Head and Neck Cancer Symptom Working Group

**Affiliations:** Department of Computational and Applied Mathematics, William Marsh Rice University, Houston, TX, USA; Department of Head and Neck Surgery, The University of Texas MD Anderson Cancer Center, Houston, TX, USA; Department of Radiation Oncology, The University of Texas MD Anderson Cancer Center, Houston, TX, USA; The University of Texas MD Anderson Cancer Center UTHealth Graduate School of Biomedical Sciences, Houston, TX, USA; Department of Radiation Oncology, University Medical Center- Groningen, Groningen, NL

## Abstract

**Purpose:** To develop a decision-making tool to choose the optimal treatment for oropharyngeal squamous cell cancer (OPSCC) patients who are eligible for primary transoral robotic surgery (TORS) and primary (chemo)radiation therapy with comparable locoregional control and survival.

**Methods:** Decision tree models were constructed to study two decision-making scenarios, 1) TORS vs. definitive radiation therapy (RT), and 2) TORS vs. definitive chemo(radiation) therapy (CRT) based on well-established objective and subjective swallowing-function instruments, MD Anderson Dysphagia Inventory (MDADI), MD Anderson Symptom Inventory–Head and Neck Module **(MDASI-HN)**, and clinician-rated Dynamic Imaging Grade of Swallowing Toxicity (**DIGEST**) that measure swallowing-related toxicity pre-therapy, 3-6 months (short-term), and 18-24 months (long-term) after therapy for five treatment cohorts (RT, CRT, TORS, TORS with adjuvant RT, and TORS with adjuvant CRT). The optimal treatment was sought as a function of postoperative extranodal extension (ENE) and/or positive margin (PM) that can trigger the adjuvant therapy. 2D heatmaps were constructed indicating the thresholds of postoperative events likelihoods required for TORS or definitive therapy to become the optimal treatment. Additionally, a risk calculation model was developed to quantify the risk associated with TORS in the settings that estimation of postoperative complications likelihoods may not be available.

**Results:** Under the first scenario and for short-term measures, MDADI and MDASI instruments indicate the superiority of definitive therapy to TORS at all times, while DIGEST required a maximum of 40% likelihood for both ENE and PM events to indicate TORS as the optimal treatment. For 18-24 months measures, MDASI indicated TORS as the optimal treatment; however, MDADI- and DIGEST-based long-term measures indicated threshold likelihoods of 90% and 25%, respectively, for TORS to remain the optimal treatment. For short-term outcomes, TORS resulted in higher toxicity even when the likelihood associated with postoperative tumor resection margin (TM) are extremely low. For higher probability of postoperative TM, all instruments indicated high risk associated with TORS (>83%). For long-term swallowing-related toxicity, TORS remained the optimal therapy independent from the probability of postoperative TM based on MDASI instrument. However, MDADI-based measure assigned a high risk to TORS (>86%) when postoperative TM is extremely likely. DIGEST-based measures indicated a very high risk associated with TORS independent from the postoperative TM likelihood (>91%).

Under the second scenario (TORS vs. definitive CRT), Both MDASI- and MDADI-based short-term measures indicated TORS as the optimal therapy independent from postoperative ENE/PM events. However, according to the DIGEST-based measure, definitive RT remained the optimal therapy when the probability of postoperative ENE and PM events exceed 80%. In this case, TORS was the optimal therapy if both events were not relatively likely (<55%). The same result was observed for MDADI and MDASI instruments for long-term measures indicating TORS as the optimal therapy independent from postoperative PM or ENE events. However, the DIGEST measure indicated that TORS is the optimal therapy only if the likelihood of both events are very low (<20%). When the postoperative TM is very unlikely (<10%), all instruments indicated TORS as the optimal therapy based on short-term outcomes. However, when the postoperative TM is extremely likely (>90%), DIGEST-based measures demonstrated the superiority of definitive CRT once the likelihood of postoperative ENE or PM is, at least, 60%. Long-term measures indicated higher sensitivity to postoperative TM likelihood with both MDADI- and DIGEST-based measures indicating a moderately high risk (> 60%) with TORS causing higher clinician-rated swallowing toxicity compared to definitive CRT when postoperative TM is extremely likely.

**Conclusion:** The current study using decision modeling shows proof of concept that in the absence of reliable estimation of postoperative ENE/PM events concurrent with significant postoperative positive margins (i.e., more than 2mm) that can trigger adjuvant therapy, the overall toxicity level incurred by OPSCC patients undergoing TORS may become more severe compared to patients receiving non-surgical treatments thus advocating definitive (C)RT protocols. The results further demonstrated that, when available, the likelihoods of postoperative events triggering postoperative adjuvant therapy must be incorporated when choosing the optimal treatment plan for eligible patients.

## 1. Introduction

Recent studies indicate that the incidence of human papillomavirus associated (HPV+) head-and-neck (HNC) cancer have been on a sharp rise, and the incidence of this malignancy is projected to nearly double by the year 2030. ^1,2^ With a yearly incidence of 600,000 cases worldwide, there are 62,000 HNC cases annually in US with an estimated 13,000 deaths ^3,4^, this is driven by the endemic rise of oropharyngeal squamous cell cancer (OPSCC) ^5^. Historical OPSCC surgical treatment for OPSCC involved open surgery such as transmandibular and transcervical pharyngotomy, which are associated with significant functional morbidity such as dysphagia. ^6–8^ To reduce postoperative morbidity, high-dose radiation therapy (RT) in combination with chemotherapy became the standard organ-preserving approach, offering comparable locoregional control and survival. However, nonsurgical (chemo) radiation therapy (CRT) treatments also put the patient at risk for multiple post-treatment toxicities, including radiation-associated dysphagia.

Transoral robotic surgery (TORS) is a surgical approach that was approved by FDA in 2009 and involves minimal disturbance to critical nerves and swallow musculature of the laryngopharynx, thus promising superior acute postoperative swallowing outcomes compared to traditional open surgical approaches ^9^ and therapeutic nonsurgical organ preservation regimens in OPSCC ^10^ as reported in prospective cohort studies. ^11–14^ Proponents of a primary TORS approach further cite potential for de-escalation protocols or avoidance of of adjuvant therapies, altogether as a major functional advantage of TORS. ^10^ Despite TORS promise, it is reported that only 9-27% of patients treated with frontline TORS avoid postoperative adjuvant RT, and 34-45% avoid adjuvant CRT. ^10^ As probabilistic outcomes of TORS, postoperative positive margins (PM) or pathological extranodal extensions (ENE) indicate increased risk of recurrence, and necessitate adjuvant (C)RT (Figure 1) with resultant short- and long-term radiation associated toxicities. ^14,15^ In a recent study on a cohort of low- and intermediate-risk OPSCC patients receiving definitive (C)RT and TORS (possibly followed by adjuvant therapy), swallowing outcomes (at 3-6 months post-treatment) were reported to be similar regardless of the primary treatment modality. ^14^ This suggests that patients undergoing TORS, even when followed by adjuvant therapy, may not incur less severe dysphagia compared to receiving definitive (chemo) radiation therapy as the primary treatment modality.

**Figure 1:**
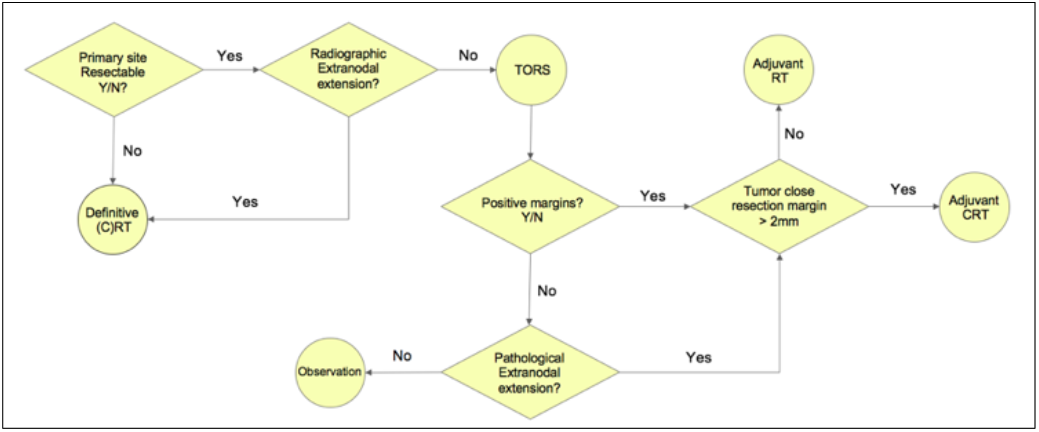
The underlying process for determining *the eligibility of the patient for TORS and its probabilistic outcomes*

At present, the relative selection criteria for surgery are primarily qualitative and subjective physician assessments. Studies suggest that, at least in current practices, physicians are quite poor at predicting the necessity of adjuvant therapy based on pre-surgical or imaging risk features. This leaves the provider and patient with an upfront pre-therapy choice: choose definitive (C)RT with *known* quantized patient-specific toxicity risk probability *OR* choose an *a priori* quantifiable toxicity risk of surgery AND an *undefined* probability of the risk toxicity of adjuvant therapy. The premise of this study is to define, the proportional likelihood of surgical risk features (and resultant indication for adjuvant therapy-associated toxicity) and? mathematically optimal decision making? between primary therapies: (chemo)RT or TORS. Put simply, we address the question of how *numerically confident* the surgeon and radiation oncologist must be in the risk of *pre-treatment* pathologic margin positivity or extranodal extension be *to rationally* select TORS for the purposes of minimizing toxicity (assuming equivalent locoregional control) [REF ORATOR, ECOG 3311].

The primary focus of the present study was to develop a decision support tool that aids in selecting the best primary treatment protocol by incorporating the likelihood of postoperative PM and/or ENE to quantify both overall therapy-related discomfort level and swallowing function impairment as short- and long-term toxicities, using an existing prospective dataset. The aims of this study were as follows: 1) to quantify the swallowing-related toxicity levels of definitive therapies (chemo RT) and TORS based on subjective and objective instruments using short-term and long-term assessments of toxicities, and 2) to determine the required confidence level of likelihood of ENE/PM to in order to determine the optimal primary therapy and its’ associated risk level. To achieve this aim, we incorporate the quantified expected value of swallowing-related outcomes of each primary treatment based on probabilistic postoperative PM and/or ENE events. This is the first application of decision analysis, a widely established tool for decision making in uncertain environments. We use this tool to quantify the risk of postoperative swallowing-related toxicities and the impact on quality of life, measured using highly reliable functional endpoints frequently used in OPSCC.

## 2. Methods

### Study Design

This secondary analysis was conducted using prospective registry data from the MD Anderson Oropharynx Cancer Registry (PA14-0947) Patient-Reported Outcomes and Function (PROF) Core. The PROF registry enrolls all consenting OPSCC/HNC patients at the University of Texas MD Anderson Cancer Center (MDACC) beginning in March 2015. The sample for this secondary analysis included patients enrolled on PA14-0914 from March 2015 to September 2019. Eligibility criteria were as follows: (1) cancer of the oropharynx, and (2) TORS or RT as primary treatment approach at MDACC. All primary treatment was determined by Multidisciplinary Tumor Board. Data analysis occurred under approval of the Institutional Review Board (protocol PA11-0809). ^14^

### Demographics

Demographics and treatment characteristics of the cohort are listed in Table 1. ^14^

**Table 1:** Characteristics of the 257 Patients With Low- to Intermediate-Risk Oropharyngeal Cancer Included in the Study ^14^

| Characteristic | All Patients<br>(N=257) | Primary<br>TORS (n=75) | Primary RT<br>(n=182) | Effect Size<br>(95% CI) |
| --- | --- | --- | --- | --- |
| Age at primary treatment start, mean (SD), y | 59.54 (9.07) | 58.70 (9.60) | 59.89 (8.84) | NA |
| Sex |  |  |  |  |
| Female | 35 (13.6) | 10 (13.3) | 25 (13.7) | NA |
| Male | 222 (86.4) | 65 (86.7) | 157 (86.3) |  |
| Primary tumor site |  |  |  |  |
| Tonsil | 135 (52.5) | 38 (50.7) | 97 (53.3) | NA |
| BOT | 116 (45.1) | 34 (45.3) | 82 (45.0) |  |
| GPS | 6 (2.3) | 3 (4.0) | 3 (1.6) |  |
| Clinical T stage of primary tumor (AJCC Staging Manual, 7th ed.) |  |  |  |  |
| 1 | 124 (48.2) | 40 (53.3) | 84 (46.2) | NA |
| 2 | 126 (49.0) | 34 (45.3) | 92 (50.6) |  |
| 3 | 7 (2.7) | 1 (1.3) | 6 (3.3) |  |
| Baseline primary tumor volume, median (range), cm<sup>3</sup> | 6.50<br>(0.30-29.30) | 5.15<br>(0.30-21.90) | 6.95<br>(0.50-29.30) | 0.18<br>(0.04-0.29) |
| Clinical N stage of primary tumor (AJCC Staging Manual, 7th ed) |  |  |  |  |
| N0 | 50 (19.5) | 31 (41.3) | 19 (10.4) | 0.40 (0.27-0.51) |
| N1 | 35 (13.6) | 15 (20.0) | 20 (11.0) |  |
| N2a | 19 (7.4) | 5 (5.3) | 15 (8.2) |  |
| N2b | 153 (59.2) | 25 (33.3) | 128 (70.3) |  |
| Induction Chemotherapy | 21 (8.2) | 4 (5.3) | 17 (9.3) | NA |
| Radiotherapy | 219 (85.2) | 37 (49.3) | 182 (100) | NA |
| Concurrent chemotherapy and radiotherapy | 162 (63.0) | 15 (20.0) | 147 (80.8) | NA |
| Radiotherapy dose, median (range), cGy | 6996<br>(6000-7000) | 6000<br>(5000-6996) | 6996<br>(6000-7000) | 0.61<br>(0.52-0.69) |
| Radiotherapy laterality |  |  |  |  |
| No radiotherapy | 38 (14.8) | 38 (50.7) | 0 | 0.67<br>(0.58-0.75) |
| Unilateral | 54 (21.0) | 17 (22.7) | 37 (20.3) |  |
| Bilateral | 165 (64.2) | 20 (26.7) | 145 (49.7) |  |
| Proton radiation therapy | 44 (20.1) | 5 (13.5) | 39 (21.4) | NA |
| Neck dissection | 93 (36.2) | 75 (100) | 18 (9.9) | 0.85<br>(0.79-0.91) |
| Baseline DIGEST score |  |  |  |  |
| 0 | 200 (80.6) | 53 (73.6) | 147 (83.3) | 0.13<br>(0.0-0.22) |
| 1 | 44 (17.7) | 16 (22.2) | 28 (16.1) |  |
| 2 | 4 (1.6) | 3 (4.2) | 1 (0.6) |  |
| 3 | 0 | 0 | 0 |  |
| Baseline MDADI global score |  |  |  |  |
| 40 | 7 (3.0) | 3 (4.6) | 4 (2.4) | NA |
| 60 | 5 (2.1) | 2 (3.0) | 3 (1.8) |  |
| 80 | 42 (17.9) | 12 (18.2) | 30 (17.6) |  |
| 100 | 181 (77.0) | 49 (74.2) | 132 (78.1) |  |
| Baseline MDADI composite score, median (range) | 95.79<br>(54.74-100) | 94.74<br>(54.74-100) | 95.79<br>(57.89-100) | NA |
| Baseline MDADI physical score, median (range) | 100 (45-100) | 100 (45-100) | 100 (55-100) | NA |
| Baseline MDADI emotional score, median (range) | 86.67<br>(43.33-100) | 86.67<br>(43.33-100) | 86.67<br>(56.67-100) | NA |
| Baseline MDADI functional score, median (range) | 100 (64-100) | 100 (68-100) | 100 (64-100) | NA |
| Baseline MDASI-HN swallowing symptom severity item score, median (range) | 0 (0-9) | 0 (0-3) | 0 (0-9) | NA |
| Baseline MDASI-HN swallowing symptom severity item score category |  |  |  |  |
| None | 133 (68.6) | 24 (72.7) | 109 (67.7) | NA |
| Mild | 56 (28.9) | 9 (27.3) | 47 (29.2) |  |
| Moderate to severe | 5 (2.6) | 0 | 5 (3.1) |  |
| Baseline MDASI-HN choke score, median (range) | 0 (0-8) | 0 (0-8) | 0 (0-7) | NA |
| Baseline MDASI-HN choke score category |  |  |  |  |
| None | 156 (80.0) | 30 (88.2) | 126 (78.3) | NA |
| Mild | 35 (18.0) | 3 (8.8) | 32 (19.9) |  |
| Moderate to severe | 4 (2.0) | 1 (2.9) | 3 (1.9) |  |

### Toxicity Measures

Prospective collection of clinician- and patient-graded outcome measures occurred at routine timepoints. The MD Anderson Dysphagia Inventory **(MDADI)** is a patient-administered 20-item questionnaire that evaluates the impact of dysphagia on quality of life. The MDADI includes one question regarding global function and 19 items that focus on the physical, emotional and functional aspects of swallowing, which are pooled and averaged to obtain a composite score (varying from 20 (poor swallowing-related quality of life) to 100 (optimal swallowing-related quality of life)) ^16^. The MD Anderson Symptom Inventory–Head and Neck Module **(MDASI-HN)** score as a validated multi-symptom inventory of patient-reported swallowing and chewing difficulties based on scores varying from 0 (symptom not present) to 10 (highest imaginable severity of the symptom) and represents a generalizable pan-symptom toxicity metric ^17^. Lastly, the Dynamic Imaging Grade of Swallowing Toxicity (**DIGEST**) as a validated and reliable objective tool that measures the presence and severity of pharyngeal dysphagia. The DIGEST parallel’s with CTCAE criteria for toxicity reporting Grade 0 (no pharyngeal dysphagia), 1 (mild), 2 (moderate), 3 (severe), and 4 (life-threatening dysphagia).^14,18^ The study was conducted using multiple instruments to avoid risk/decision calibration predicated only based on a subset of the patient toxicity profile, thus accounting for inter-therapy differential toxicity.

### Measures

A total of six measures were developed for this study as follows: each instrument was assessed for all treatment cohorts (TORS, RT alone, CRT alone, TORS with adjuvant RT (TORS+RT), TORS with adjuvant CRT (TORS+CRT)) pre-therapy (baseline), 3-6 months and 18-24 months after primary treatment (Table 2). For each cohort, MDADI-based and MDASI-based *absolute short-term deterioration* in swallowing function (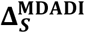 and 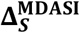) were defined as the reduction in MDADI baseline score and the increment in MDASI baseline score, respectively, within 3-6 months. MDADI-based and MDASI-based *absolute long-term deterioration* in swallowing function (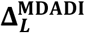 and 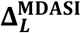) were defined analogously with respect to 18-24 months scores. For each treatment cohort, DIGEST-based *short- and long-term deteriorations* in swallowing functions (***D***^**DIGEST**^ and ***R***^**DIGEST**^) were calculated as the fraction of baseline population whose DIGEST baseline grades evolved into any worse grade within 3-6 and 18-24 months after receiving therapy, respectively. (Online Supplements A1-A3).

**Table 2:** MDADI, MDASI, and DIGEST pre- and post-bootstrapping scores pre-therapy (baseline), within 3-6 months, and within 18-24 months post-therapy; for DIGEST, total number of cases (**bold**) and total number of cases whose baseline grades have evolved into inferior grades.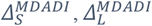: MDADI-based absolute short- and long-term deterioration;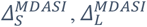: MDASI-based absolute short- and long-term deterioration; *D*^*DIGEST*^, *R*^*DIGEST*^: DIGEST-based absolute short- and long-term deterioration in swallowing function.

| Instrument | CRT | RT | TORS+<br>CRT | TORS+<br>RT | TORS |
| --- | --- | --- | --- | --- | --- |
| **MDADI** |  |  |  |  |  |
| Baseline (mean) | 93.26 | 87.17 | 94.04 | 90.18 | 88.69 |
| 3-6 Months | 81.14 | 82.02 | 82.89 | 83.80 | 82.55 |
| 18-24 Months | 86.56 | 81.98 | 85.16 | 88.28 | 86.05 |
| Absolute short-term deterioration $\Delta_S^{MDADI}$ (Bootstrapped) | 12.1 | 5.34 | 11.27 | 6.43 | 6.3 |
| Absolute long-term deterioration $\Delta_L^{MDADI}$ (Bootstrapped) | 6.71 | 5.25 | 8.81 | 2.05 | 2.57 |
| **MDASI** |  |  |  |  |  |
| Baseline (mean) | 0.47 | 0.99 | 0.59 | 0.32 | 0.81 |
| 3-6 Months | 1.43 | 1.37 | 1.09 | 0.91 | 1.20 |
| 18-24 Months | 0.99 | 1.35 | 0.62 | 0.54 | 0.93 |
| Absolute short-term deterioration $\Delta_S^{MDADI}$ (Bootstrapped) | 0.95 | 0.38 | 0.51 | 0.59 | 0.39 |
| Absolute long-term deterioration $\Delta_L^{MDADI}$ (Bootstrapped) | 0.51 | 0.36 | 0.05 | 0.22 | 0.12 |
| **DIGEST** |  |  |  |  |  |
| 3-6 Months incidence with worsen grade | 60 (**120**)* | 10 (**23**) | 4 (**10**) | 11 (**16**) | 5 (**24**) |
| 18-24 Months incidence with worsen grade | 23 (**66**) | 3 (**11**) | 3 (**6**) | 7 (**7**) | 1 (**14**) |
| Absolute short-term deterioration $D^{DIGEST}$ (Bootstrapped) | 0.5 | 0.44 | 0.4 | 0.69 | 0.21 |
| Absolute long-term deterioration $R^{DIGEST}$ (Bootstrapped) | 0.35 | 0.27 | 0.5 | 1.00 | 0.07 |

\*: indicates number of patients with worsen condition out of all patients.

### Statistical Analysis

This study was based on a published analysis conducted by Hutcheson et al. ^14^ using the same patient cohort. Further, for each treatment cohort, bootstrapping-based resampling ^19^ was applied to mitigate the effects of unequal sample sizes across treatment cohorts (n=10,000). For MDADI and MDASI scores, bootstrapping was employed based on the empirical distribution computed from the frequency of observed scores. The values of 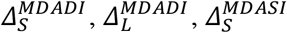 and 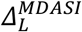 were calculated using bootstrapped datasets for each treatment cohort. For DIGEST grades, bootstrapping was employed based on the assumption that the evolution of baseline grades into 3-6 months and 18-24 months grades follow multinomial distribution based on the incidence reported in Table C1 (Online Supplement C). Using the bootstrapped dataset, the values of *R*^*DIGEST*^ and *D*^*DIGEST*^ were calculated. Table 2 provides the bootstrap-generated measures.

### Decision Tree Analysis

The aim of this study is to seek the confidence level suggested the clinical team abide by with respect to the likelihood of postoperative ENE/PM for TORS (definitive (C)RT) to suggest the optimal primary treatment, i.e., to outperform definitive (C)RT (TORS) in terms of *expected* swallowing-related toxicity level. An expected-value decision tree was constructed following the clinical flow depicted in Figure 1 allowing the measure-based comparison of definitive (C)RT having deterministic outcomes and TORS having probabilistic outcomes (Figure 2). As one of the most common decision support tools for medical decision making, decision trees are extremely efficient in terms of implementing medical guidelines for scenarios with probabilistic outcomes to determine the optimal decision based on of expected value of the outcomes. ^20^

**Figure 2:**
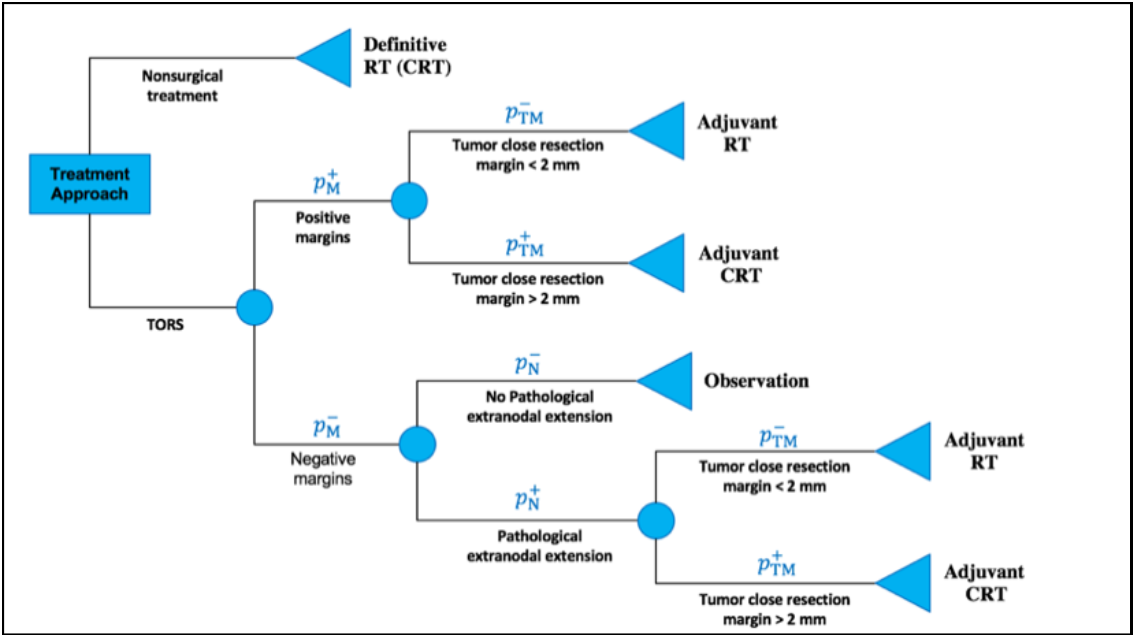
General decision Tree for TORS decision making; scenario 1: definitive RT vs. TORS; scenario 2: CRT vs. TORS; 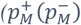: probability of having (not having) positive margins after surgery; 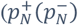: probability of having (not having) extranodal extension after surgery; 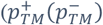: probability of having tumor resection margin of more (less) than 2 mm after surgery

The decision tree model was constructed under two distinct scenarios, 1) TORS vs. definitive RT, and 2) TORS vs. definitive CRT, based on the assumption that the patients in each scenario are eligible for both surgical and definitive therapies with comparable locoregional control and survival. For each scenario, the decision model was analyzed for each measure: using the collected measure values (Table 2), the expected value of TORS was calculated as a function of postoperative ENE and PM likelihoods (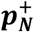 and 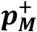, respectively, ranging from 0 to 1) based on the assumption that when an adjuvant therapy is required, it is equally likely that the patient will undergo adjuvant RT or adjuvant CRT. (Sensitivity analysis was performed to study the effects of this assumption as reported in Online Supplement B.)

In each scenario, the optimal choice between TORS and the definitive therapy was made based on the observation that for both short- and long-term swallowing-related toxicity levels, the treatment protocol having lower expected value is more favorable. For each scenario and for each short-or long-term measure, the *cut-off* value for TORS (***c***_***S***_, ***c***_***L***_) was computed as the highest possible expected measure value of TORS for which TORS remains the optimal treatment.

The results of decision tree analysis were demonstrated as 2D heatmaps, for each measure, revealing the combination of probability values for postoperative ENE and PM events for which TORS results in lower swallowing-related toxicity, thus becoming the optimal treatment. The heatmaps were also employed to derive individual postoperative ENE and PM likelihoods for which definitive therapy becomes the optimal treatment having lower swallowing-related toxicity level. Finally, to account for the inherent difficulty in pre-therapy estimation of postoperative ENE/PM likelihoods, measure-based *risk associated with* TORS (***r***) were developed as the fraction of possible combination of postoperative ENE and PM likelihoods for which definitive therapy becomes the optimal choice compared to TORS.

## 3. Results

### 3.1. TORS vs. definitive RT (*Scenario 1)*

#### 3.1.1. Short-term (3-6 month) outcomes

The decision tree analysis using both MDADI- and MDASI-based measures implied that definitive RT remains the optimal treatment protocol for any postoperative ENE and PM likelihoods (Figure 3). As evidenced by measure values in Table 2, this is justified based on the observation that the short-term MDADI- and MDASI-based swallowing-related toxicities of TORS (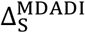 and 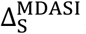, respectively) were always more severe compared to definitive RT. According to the DIGEST-measure (presence and severity of dysphagia), if the likelihood associated with either ENE or PM is, at least, 75%, definitive RT remains the optimal treatment. For cases in which the likelihood of neither ENE or PM is more than 40%, TORS was the optimal treatment. Further, in the absence of pre-therapy details regarding ENE or PM likelihood, TORS risk level was at least 65% (according to DIGEST), and surgery was not recommended based on MDADI- and MDASI-based measures.

**Figure 3:**
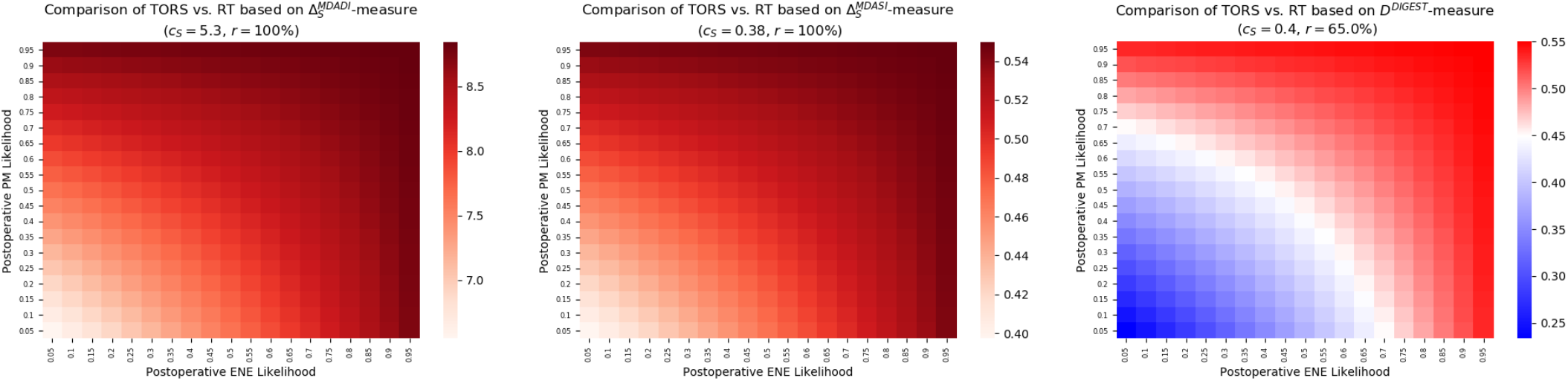
Expected deterioration in swallowing function due to TORS and definitive RT based on short-term measures (left) MDADI, (center) MDASI, (right) DIGEST.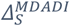: MDADI-based absolute short-term deterioration;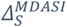: MDASI-based absolute short-term deterioration; *D*^*DIGEST*^: DIGEST-based absolute short-term deterioration in swallowing function; *c*_*S*_: cut-off value for TORS; and *r*: risk associated with TORS.

#### 3.1.2. Long-term (18-24 month) outcomes

For long-term measures, TORS was the optimal treatment based on MDASI instrument for any confidence level regarding the likelihood of postoperative ENE and PM (Figure 4). However, based on MDADI instrument, definitive RT became the optimal treatment when either postoperative ENE or PM are extremely likely (> 90%) to occur. TORS was the optimal treatment if the likelihood of both postoperative events remained less than 70%. According to DIGEST instrument, however, definitive RT remained the optimal treatment even if either of the events was likely, at least, 25%. In this case, TORS becomes the optimal treatment plan only if both events are extremely unlikely to occur (<10%). Finally, in the absence of pre-therapy information about ENE or PM likelihood, TORS risk level is at 21% according to MDADI, and 97% according to DIGEST with TORS carrying no risk based on MDASI instrument.

**Figure 4:**
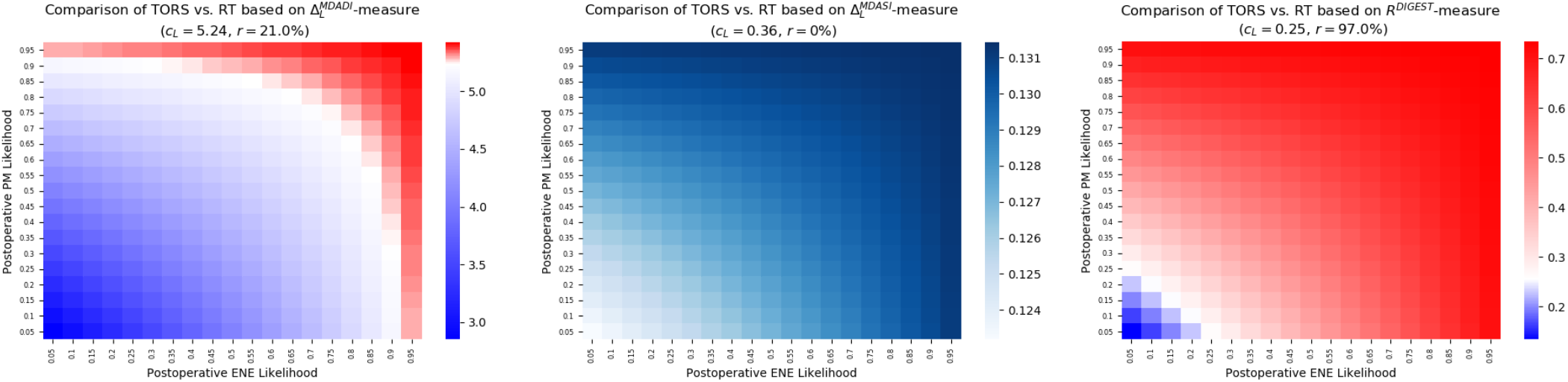
Expected deterioration in swallowing function due to TORS and definitive RT based on long-term measures (left) MDADI, (center) MDASI, (right) DIGEST.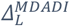: MDADI-based absolute short-term deterioration;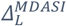: MDASI-based absolute short-term deterioration; R^DIGEST^: DIGEST-based absolute short-term deterioration in swallowing function; *c*_*S*_: cut-off value for TORS; and r: risk associated with TORS.

Table 3 summarizes the confidence level of postoperative events likelihood required to ensure TORS or definitive RT is the optimal treatment under the first scenario.

**Table 3:**
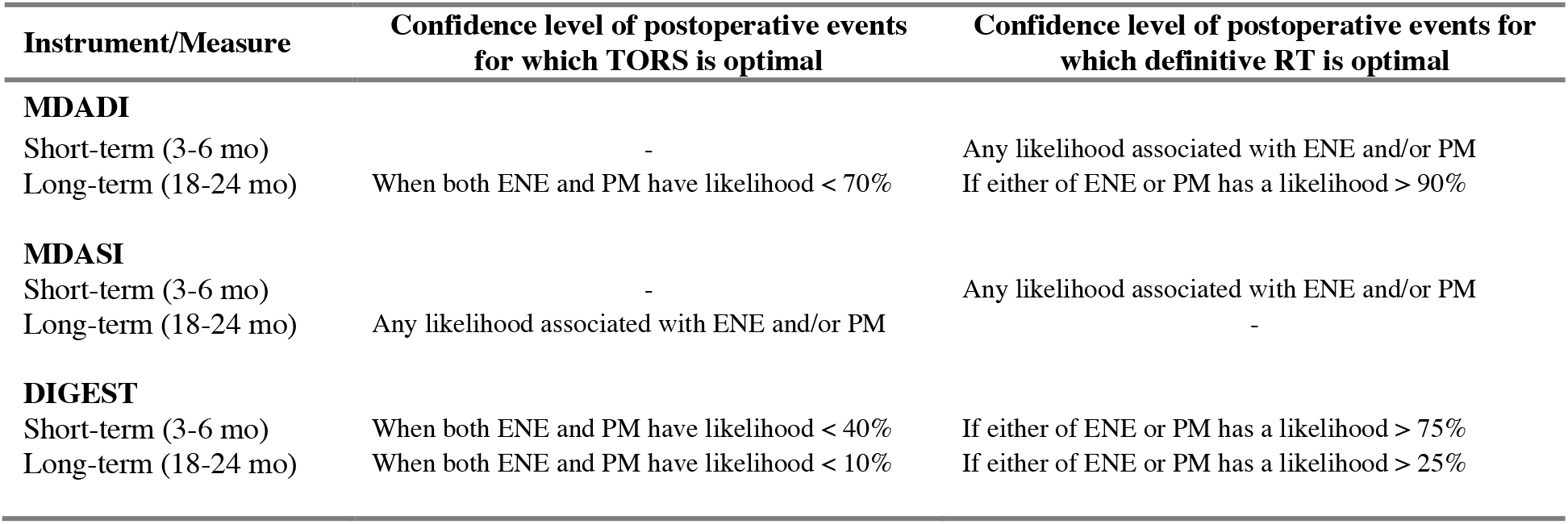
Range of likelihoods required for TORS or definitive RT to become the optimal treatment under the first scenario. ENE: postoperative extranodal extension; PM: postoperative positive margin.

| Instrument/Measure | Confidence level of postoperative events for which TORS is optimal | Confidence level of postoperative events for which definitive RT is optimal |
| --- | --- | --- |
| **MDADI** |  |  |
| Short-term (3-6 mo) | - | Any likelihood associated with ENE and/or PM |
| Long-term (18-24 mo) | When both ENE and PM have likelihood < 70% | If either of ENE or PM has a likelihood > 90% |
| **MDASI** |  |  |
| Short-term (3-6 mo) | - | Any likelihood associated with ENE and/or PM |
| Long-term (18-24 mo) | Any likelihood associated with ENE and/or PM | - |
| **DIGEST** |  |  |
| Short-term (3-6 mo) | When both ENE and PM have likelihood < 40% | If either of ENE or PM has a likelihood > 75% |
| Long-term (18-24 mo) | When both ENE and PM have likelihood < 10% | If either of ENE or PM has a likelihood > 25% |

### 3.2. TORS vs. definitive CRT *(Scenario 2)*

#### 3.2.1. Short-term (3-6 month) outcomes

When comparing TORS to definitive CRT using short-term measures, TORS was the optimal treatment based on both MDADI and MDASI instruments for any likelihoods associated with postoperative ENE and/or PM (Figure 5). This observation was evident from the related measure values reported in Table 2. Based on DIGEST instrument, the patient undergoing TORS will experience higher swallowing-related toxicity if the likelihood for any of the postoperative event is more than 80%, thus making definitive CRT the optimal treatment in those cases. TORS remained the optimal treatment when both postoperative events have a likelihood of, at most, 55%. Further, in the absence of pre-therapy information about ENE or PM likelihood, TORS risk level is at most 45% according to DIGEST, while it carries no risk according to the other instruments.

**Figure 5:**
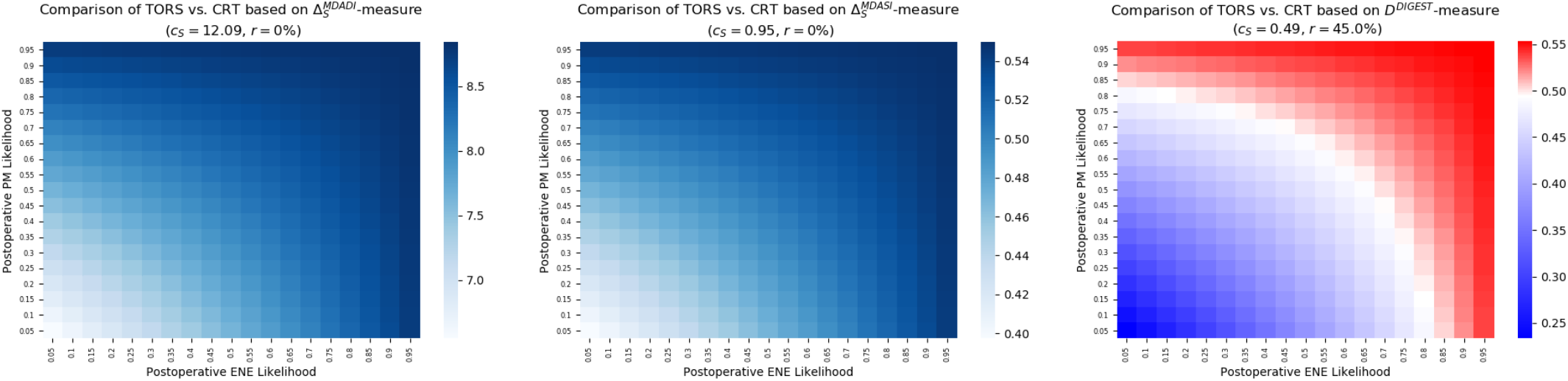
Expected deterioration in swallowing function due to TORS and definitive CRT based on short-term measures (left) MDADI, (center) MDASI, (right) DIGEST.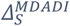: MDADI-based absolute short-term deterioration; 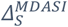: MDASI-based absolute short-term deterioration; *D*^*DIGEST*^: DIGEST-based absolute short-term deterioration in swallowing function; *c*_*S*_: cut-off value for TORS; and *r*: risk associated with TORS.

#### 3.2.2. Long-term (18-24 month) outcomes

For this scenario, the results of decision tree analysis for long-term measures were highly similar to those for the first scenario (Figure 6). In particular, TORS remained the optimal treatment based on both MDADI and MDASI instruments being insensitive to the likelihood of postoperative ENE or PM. However, according to DIGEST instrument, even moderate likelihood (>40%) for either of postoperative events implies the superiority of definitive CRT over TORS. The latter becomes the optimal treatment when both postoperative events have a small likelihood (<20%). Further, when no pre-therapy information about the likelihoods is available, DIGEST assigns a risk level of 91% to TORS, while MDASI and MDASI indicate that there is no risk level for TORS (Figure 6).

**Figure 6:**
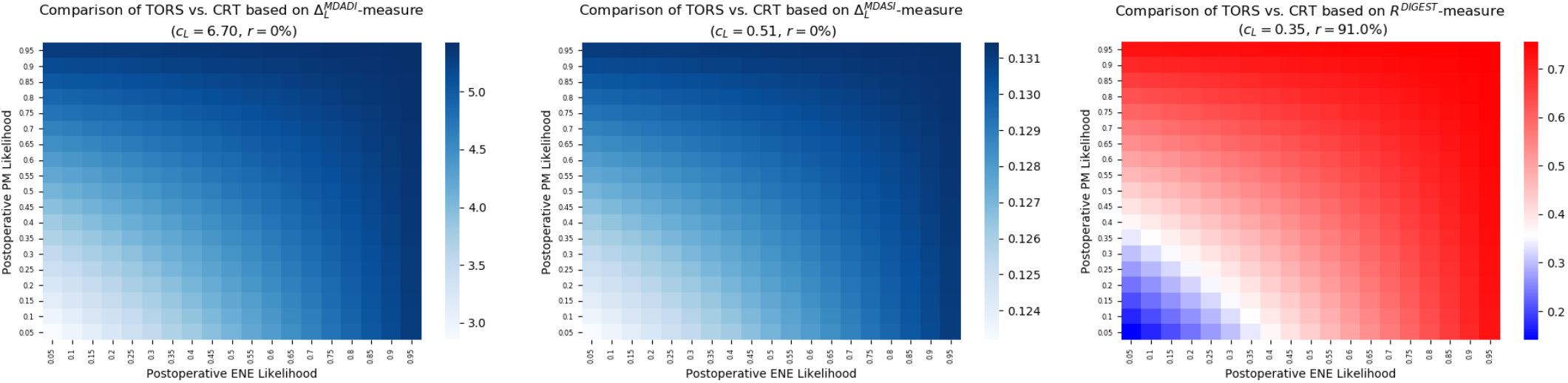
Expected deterioration in swallowing function due to TORS and definitive CRT based on long-term measures (left) MDADI, (center) MDASI, (right) DIGEST.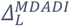: MDADI-based absolute short-term deterioration;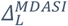: MDASI-based absolute short-term deterioration; *R*^*DIGEST*^: DIGEST-based absolute short-term deterioration in swallowing function; *c*_*S*_: cut-off value for TORS; and *r*: risk associated with TORS.

A summary of the confidence level of postoperative events likelihoods required for the optimality of TORS and definitive CRT is given in Table 4.

**Table 4:** Range of likelihoods required for TORS or definitive CRT to become the optimal treatment under the second scenario. ENE: postoperative extranodal extension; PM: postoperative positive margin.

| Instrument/Measure | Confidence level of postoperative events for which TORS is optimal | Confidence level of postoperative events for which definitive CRT is optimal |
| --- | --- | --- |
| **MDADI** |  |  |
| Short-term (3-6 mo) | Any likelihood associated with ENE and/or PM | - |
| Long-term (18-24 mo) | Any likelihood associated with ENE and/or PM | - |
| **MDASI** |  |  |
| Short-term (3-6 mo) | Any likelihood associated with ENE and/or PM | - |
| Long-term (18-24 mo) | Any likelihood associated with ENE and/or PM | - |
| **DIGEST** |  |  |
| Short-term (3-6 mo) | When both ENE and PM have likelihood $< 55\%$ | If either of ENE or PM has a likelihood $> 80\%$ |
| Long-term (18-24 mo) | When both ENE and PM have likelihood $< 20\%$ | If either of ENE or PM has a likelihood $> 40\%$ |

## 4. Discussion

Since the approval of TORS by FDA as a minimally invasive surgical treatment protocol for HNC patients, there have been several studies reporting on the success of TORS as an option for treatment of early-stage oropharyngeal carcinomas due to its favorable oncologic outcomes and its potential to mitigate the toxicities incurred by patients in other surgical techniques or primary (chemo) radiation therapy. ^21–24^ Consequently, TORS has been increasingly used for low- to intermediate-risk OPSCC patients having small-volume primary tumor and near-normal baseline function ^14^. However, studies suggest a considerable percentage of patients that have undergone postoperative adjuvant (chemo) radiation therapy ^25–27^, despite being theoretically believed to be candidate for surgical therapy alone. Put simply, the current data suggest that surgeons and radiation oncologists are decidedly poor at predicting whether a patient will require adjuvant treatment from pre-therapy exam, and thus many patients offered surgical resection are in fact being offered *not* TORS alone, but *rather some unquantified probability of double- or triple-modality therapy* (and the concomitant additional toxicities therefrom). This is primarily due to absence of extreme *pre*-surgical certitude regarding of *post*-TORS histopathological features, which makes it a challenging decision-making problem to choose between initial TORS or definitive non-surgical treatment protocols.

Operations research is an applied mathematics discipline focused on optimization of decision functions by rigorous statistical modeling. As one of the widely established toolsets in Operations Research, decision analysis provides an integrated framework to study decision-making scenarios that involve uncertain outcomes. The decision analysis model developed in this study incorporates the imputed pre-therapy physic-assessed statistical likelihood of the two major postoperative indicator events that trigger adjuvant therapy; namely *pre-therapy* physician-estimated probability of margin positivity and extranodal extension. Through quantifying *post-therapy* swallowing-related toxicities using well-established patient-reported and objective instruments, the model in this study captures the expected outcome of a treatment regimen which in comparison to the definitive radiotherapy’s outcome can aid the clinical team in choosing the optimal treatment protocol. The results of this model can also be utilized to compute the risk level associated with TORS in terms of developing swallowing-related toxicities higher than definitive (chemo) radiation therapy in the absence of pre-therapy information on the likelihood of postoperative events that can trigger the need for adjuvant therapy.

The decision analysis model in this work has its own limitations. It currently relies on a single institutional database with a cohort of 257 patients. Bootstrapping was employed to mitigate the effects of small-size population allowing the model to make assumptions as “real-life” as possible. Further, expected-value decision analysis has its own disadvantages, namely the sensitivity to the probability values as well as measures. Sensitivity analysis was performed to determine the variation of TORS risk level as a function of the likelihood of postoperative events as well as associated quantified short-term and long-term toxicity of all treatment protocols.

Three observations are notable from the current analysis: 1) there are distinctly different optimum choices based on the probability of toxicity that differ whether radiotherapy-alone or chemoradiotherapy is the comparator for surgical treatment; 2) there are divergent optimum choice of therapy regarding subjective multi-symptom (MDASI), subjective swallowing (MDADI) or objective swallowing (DIGEST) is the toxicity metric of interest; 3) choice of therapy based on early (3-6 month) swallowing outcomes may not reflect the optimum therapy selection for later time-points (18-24 months).

Ultimately, the aim of this effort is to quantize decision making for HNC/OPSCC patients eligible for alternative treatment protocols. The vast majority of cases selected for definitive organ-reservation or surgical therapy (potentially followed by adjuvant radiotherapy) are typically made using heuristic physician-decision processes which appear to speciously high estimates of the potential for single-modality surgery. However, if advanced approaches such as improved standardized radiologic assessment ^28^, AI-assisted imaging analysis, or risk-models ^29^ could improve outcomes by bringing quantitative decision-support to surgeons and radiation oncologists. Further, this data serves to define preoperative assessment tools for decision support for future explorations.

## 5. Conclusion

Our models demonstrated optimal decision thresholds for selection of surgical +/- adjuvant therapy or organ preservation with (chemo)radiotherapy based on clinically-representative subjective and objective toxicity outcomes. The resultant thresholds for physician certainty for prediction of clinical risk features necessitating adjuvant therapy should be considered with these decision tools as a component of multi-disciplinary patient-centric therapy selection for early-stage oropharyngeal cancer patients.

## Data Availability

Anonymized data is available upon request to Kate Hutcheson at.

## Appendix A1: Computation of MDADI-based short-term and long-term scores

Based on the responses from the patients treated via treatment approach X (i.e., TORS, D_RT, D_CRT, TORS+RT, TORS+CRT, as stated in *Table 1*, the *absolute* short- and long-term reductions in MDADI score were computed as

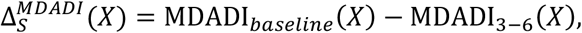

and

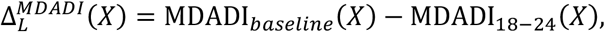

respectively, where MDADI_*baseline*_ (*X*), MDADI_3-6_(*X*), and MDADI_12-24_(*X*) are the patient-reported MDADI scores prior to receiving treatment *X*, within 3-6 months, and within 18-24 months after receiving treatment *X*, respectively, for a given treatment *X*.

## Appendix A2: Computation of MDASI-based short-term and long-term scores

Participating patients completed the MDASI-HN questionnaire before (baseline), within 3-6 month, and 18-24 months after therapy. The MDASI-based *absolute* short- and long-term increments for treatment approach X was computed as

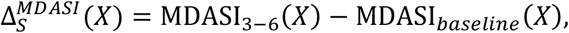

and

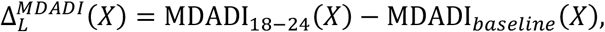

respectively.

## Appendix A3: Computation of DIGEST-based short-term and long-term grades

In this study, measures based on pre- and post-therapy DIGEST grades were employed. First, the *DIGEST-based relative increment* in the swallowing-related symptoms was defined as

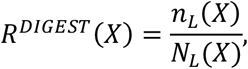

where *n*_L_(*X*) is the number of patients whose DIGEST grade increased within 18-24 months compared to their baseline grades. Also, *N*_L_(*X*) is the number of patients for whom DIGEST grade was computed within 18-24 months after the therapy. Furthermore, the *DIGEST-based relative short-term discomfort* of treatment *X* was defined as

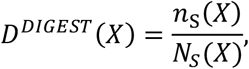

with *n*_S_(*X*) being the number of patients whose DIGEST grade increased within 3-6 months compared to their baseline grades. N_S_(*X*) is defined analogously to N_L_(*X*) for the period of 3-6 months after treatment *X*.

## Appendix B: Sensitivity analysis for the likelihood of having significant postoperative tumor resection margin

The heatmaps were also reproduced for extremely low and high values of the probability of having significant postoperative tumor resection margin 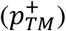 in order to study its effect on the TORS risk level compared to definitive therapies.

### B1. TORS vs. definitive RT

#### B1.1. Short-term outcomes 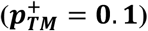

**Figure B1:**
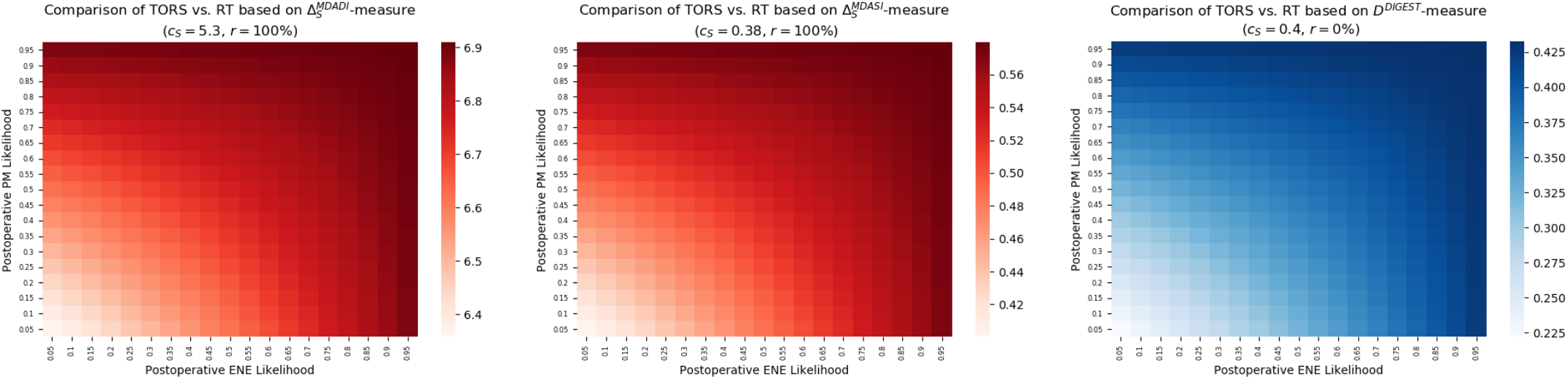
Expected deterioration in swallowing function due to TORS and definitive RT based on short-term measures (left) MDADI, (center) MDASI, (right) DIGEST.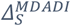: MDADI-based absolute short-term deterioration;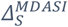: MDASI-based absolute short-term deterioration; *D*^*DIGEST*^: DIGEST-based absolute short-term deterioration in swallowing function; *c*_*S*_: cut-off value for TORS; and *r*: risk associated with TORS for 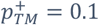.

#### B1.2. Short-term outcomes 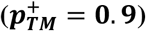

**Figure B2:**
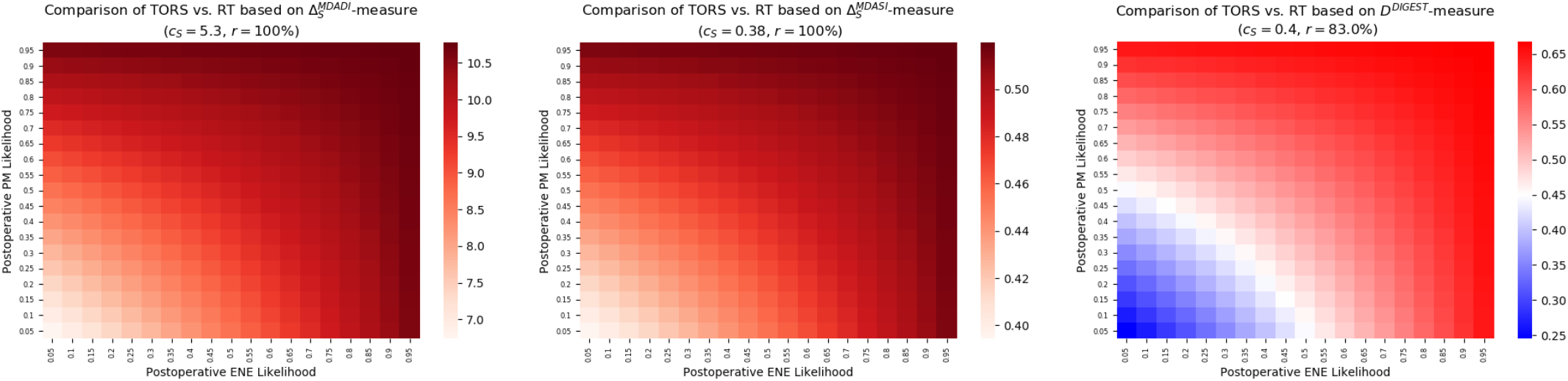
Expected deterioration in swallowing function due to TORS and definitive RT based on short-term measures (left) MDADI, (center) MDASI, (right) DIGEST.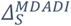: MDADI-based absolute short-term deterioration;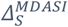: MDASI-based absolute short-term deterioration; *D*^*DIGEST*^: DIGEST-based absolute short-term deterioration in swallowing function; *c*_*S*_: cut-off value for TORS; and *r*: risk associated with TORS for 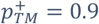.

#### B1.3. Long-term outcomes 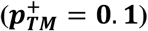

**Figure B3:**
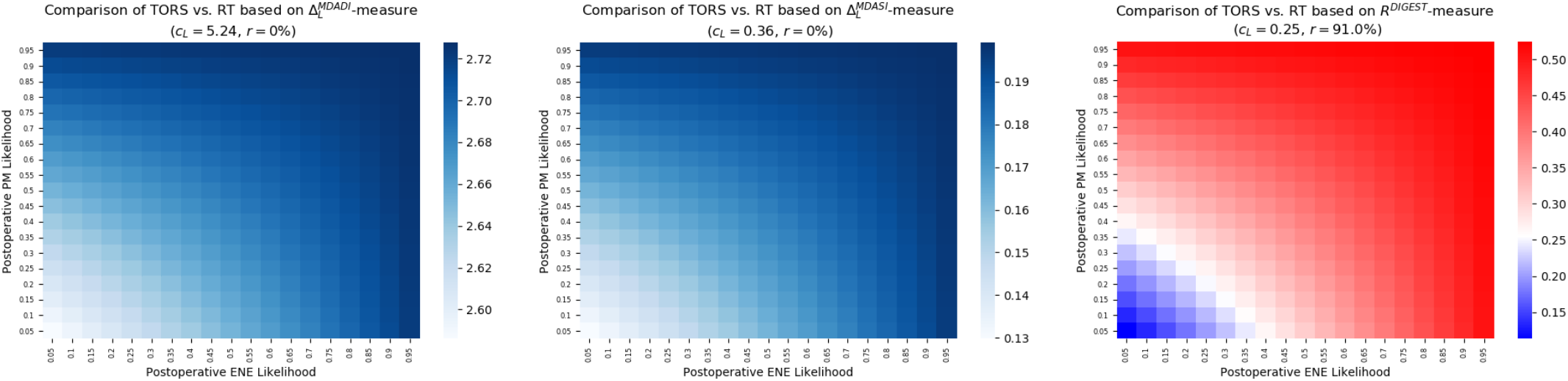
Expected deterioration in swallowing function due to TORS and definitive RT based on long-term measures (left) MDADI, (center) MDASI, (right) DIGEST.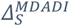: MDADI-based absolute short-term deterioration;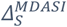: MDASI-based absolute short-term deterioration; *D*^*DIGEST*^: DIGEST-based absolute short-term deterioration in swallowing function; *c*_*S*_: cut-off value for TORS; and *r*: risk associated with TORS for 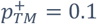.

#### B1.4. Long-term outcomes 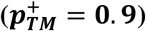

**Figure B4:**
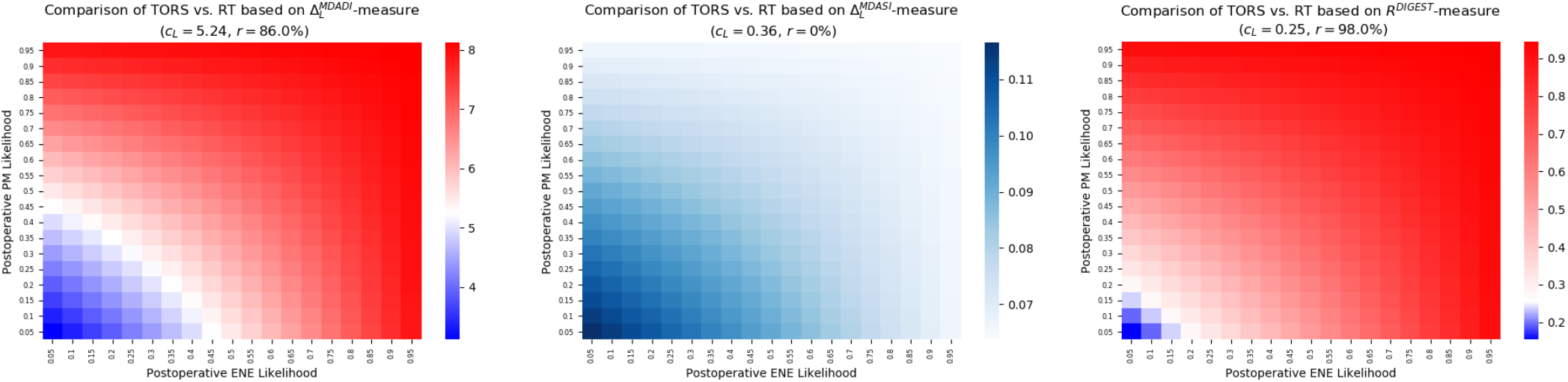
Expected deterioration in swallowing function due to TORS and definitive RT based on short-term measures (left) MDADI, (center) MDASI, (right) DIGEST. 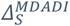: MDADI-based absolute short-term deterioration;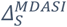: MDASI-based absolute short-term deterioration; *D*^*DIGEST*^: DIGEST-based absolute short-term deterioration in swallowing function; *c*_*S*_: cut-off value for TORS; and *r*: risk associated with TORS for 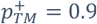.

Table B1 summarizes the risk level variation associated with TORS (when compared to definitive RT), as a function of tumor resection margin, in the absence of pre-therapy information about the postoperative ENE and PM likelihoods.

**Table B1:** Sensitivity of risk level associated with TORS (vs. definitive RT) as a function of postoperative tumor resection margin. 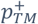: probability of having tumor resection margin > 2mm; *r*: TORS risk level; (I): Insensitive to 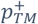 ; (HS): Highly sensitive to 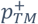 ;; (LS): Low sensitive to 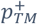.

| | $p_{TM}^+ = 50\%$ | Risk sensitivity to $p_{TM}^+$ | |
| --- | --- | --- | --- |
| Short-term toxicity level | 65% $\leq r \leq 100\%$<br>TORS carries high risk<br>(conclusive across all instruments) | MDADI | $r = 100\%$ (I) |
| | | MDASI | $r = 100\%$ (I) |
| | | DIGEST | $r \leq 83\%$ (HS) |
| Long-term toxicity level | 0% $\leq r \leq 97\%$<br>TORS risk varies based on the instrument<br>(inconclusive across all instruments) | MDADI | $r \leq 86\%$ (HS) |
| | | MDASI | $r = 0\%$ (I) |
| | | DIGEST | 91% $\leq r \leq 98\%$ (I) |

### B2. TORS vs. definitive CRT

#### B2.1. Short-term outcomes 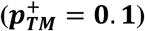

**Figure B5:**
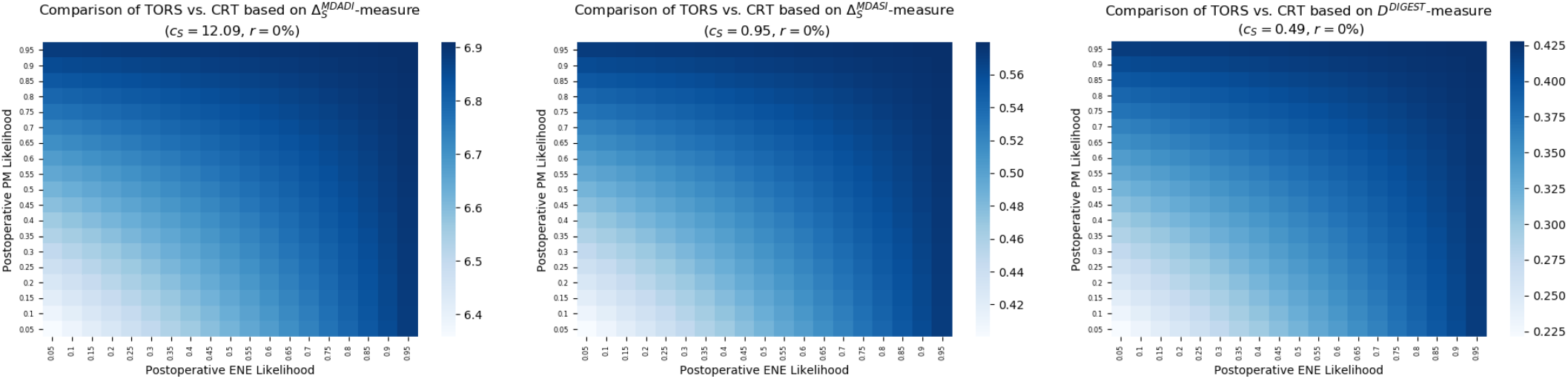
Expected deterioration in swallowing function due to TORS and definitive CRT based on short-term measures (left) MDADI, (center) MDASI, (right) DIGEST. 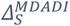: MDADI-based absolute short-term deterioration; 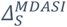: MDASI-based absolute short-term deterioration; *D*^*DIGEST*^: DIGEST-based absolute short-term deterioration in swallowing function; *c*_*S*_: cut-off value for TORS; and *r*: risk associated with TORS for 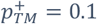.

#### B2.2. Short-term outcomes 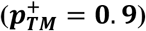

**Figure B6:**
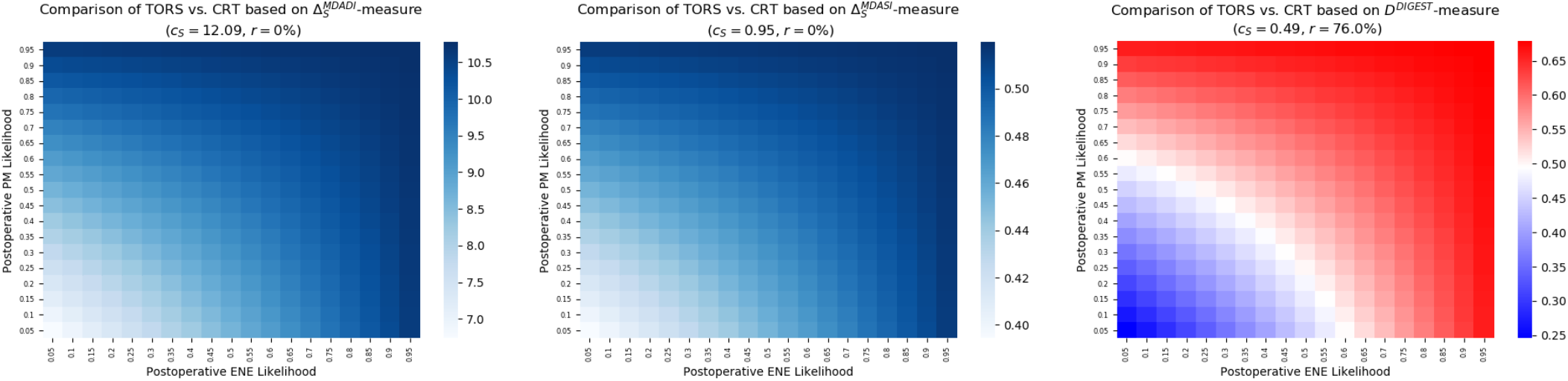
Expected deterioration in swallowing function due to TORS and definitive CRT based on short-term measures (left) MDADI, (center) MDASI, (right) DIGEST. 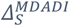: MDADI-based absolute short-term deterioration; 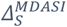: MDASI-based absolute short-term deterioration; *D*^*DIGEST*^: DIGEST-based absolute short-term deterioration in swallowing function; *c*_*S*_ cut-off value for TORS; and *r*: risk associated with TORS for 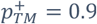.

#### B2.3. Long-term outcomes 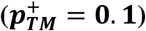

**Figure B7:**
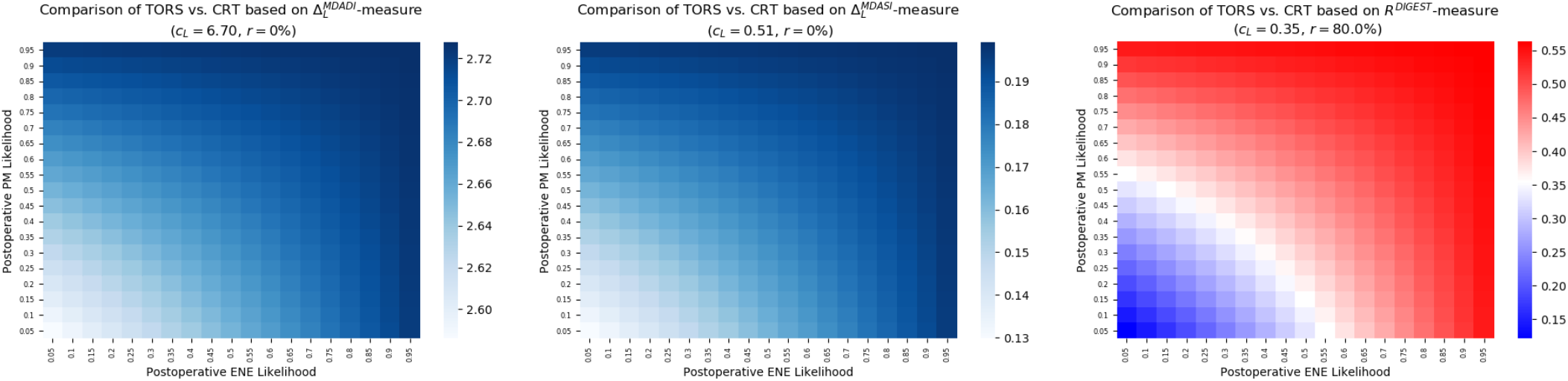
Expected deterioration in swallowing function due to TORS and definitive CRT based on long-term measures (left) MDADI, (center) MDASI, (right) DIGEST. 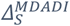: MDADI-based absolute short-term deterioration; 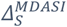: MDASI-based absolute short-term deterioration; *D*^*DIGEST*^: DIGEST-based absolute short-term deterioration in swallowing function; *c*_*S*_: cut-off value for TORS; and *r*: risk associated with TORS for 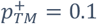.

#### B2.4. Long-term outcomes 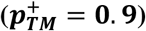

**Figure B8:**
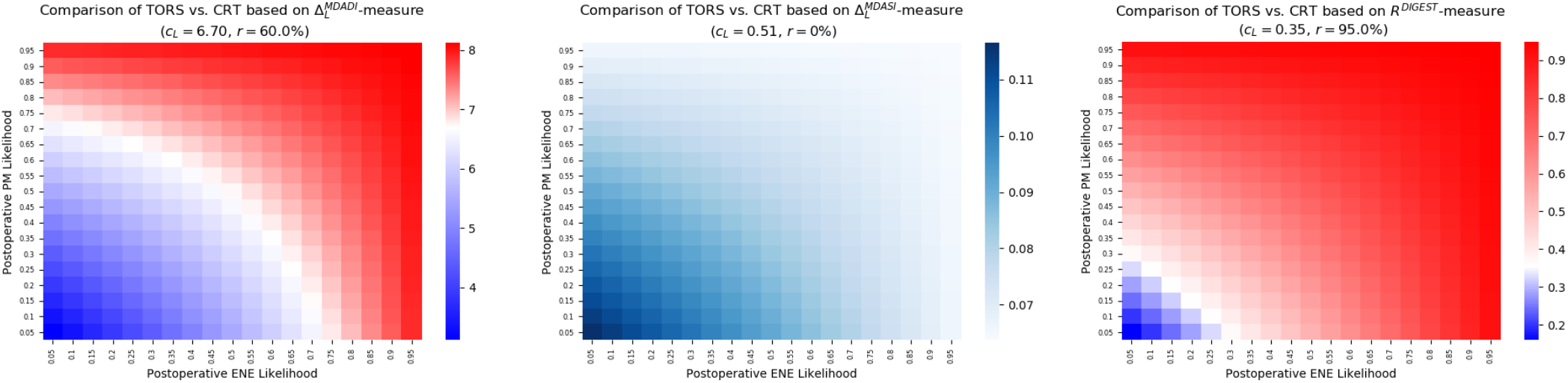
Expected deterioration in swallowing function due to TORS and definitive CRT based on long-term measures (left) MDADI, (center) MDASI, (right) DIGEST. 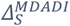: MDADI-based absolute short-term deterioration; 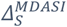: MDASI-based absolute short-term deterioration; *D*^*DIGEST*^: DIGEST-based absolute short-term deterioration in swallowing function; *c*_*S*_: cut-off value for TORS; and *r*: risk associated with TORS for 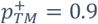.

Table B2 summarizes the risk level variation associated with TORS (when compared to definitive CRT), as a function of tumor resection margin, in the absence of pre-therapy information about the postoperative ENE and PM likelihoods.

**Table B2:**
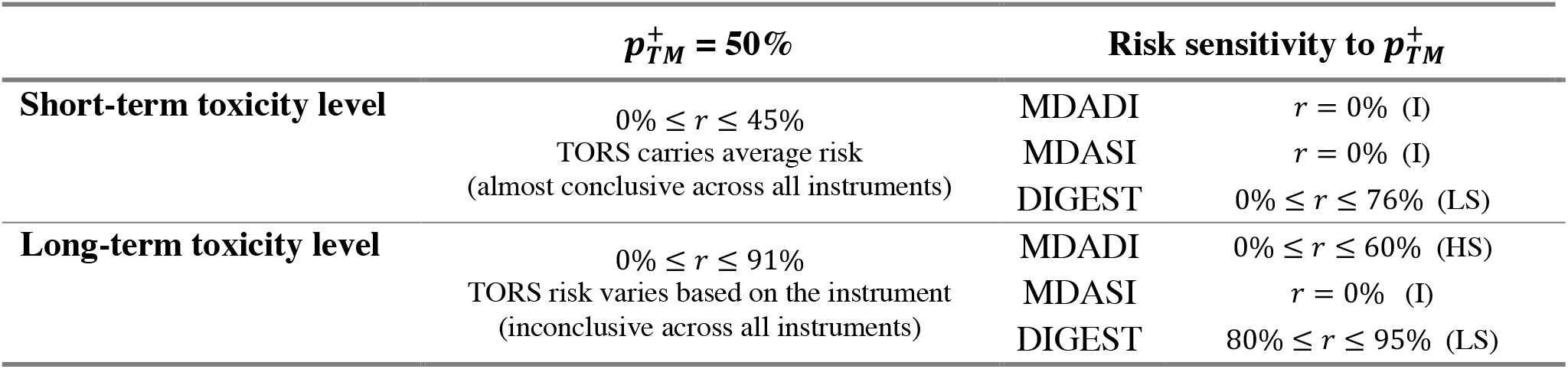
Sensitivity of risk level associated with TORS (vs. definitive CRT) as a function of postoperative tumor resection margin. 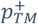 : probability of having tumor resection margin > 2mm; *r*: TORS risk level; (I): Insensitive to 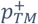; (HS): Highly sensitive to 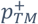 ; (RS) Relatively sensitive to 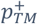; (LS): Low sensitive to 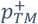;

## Appendix C: DIGEST baseline grades evolution within 3-6 and 18-24 months

**Table C1:**
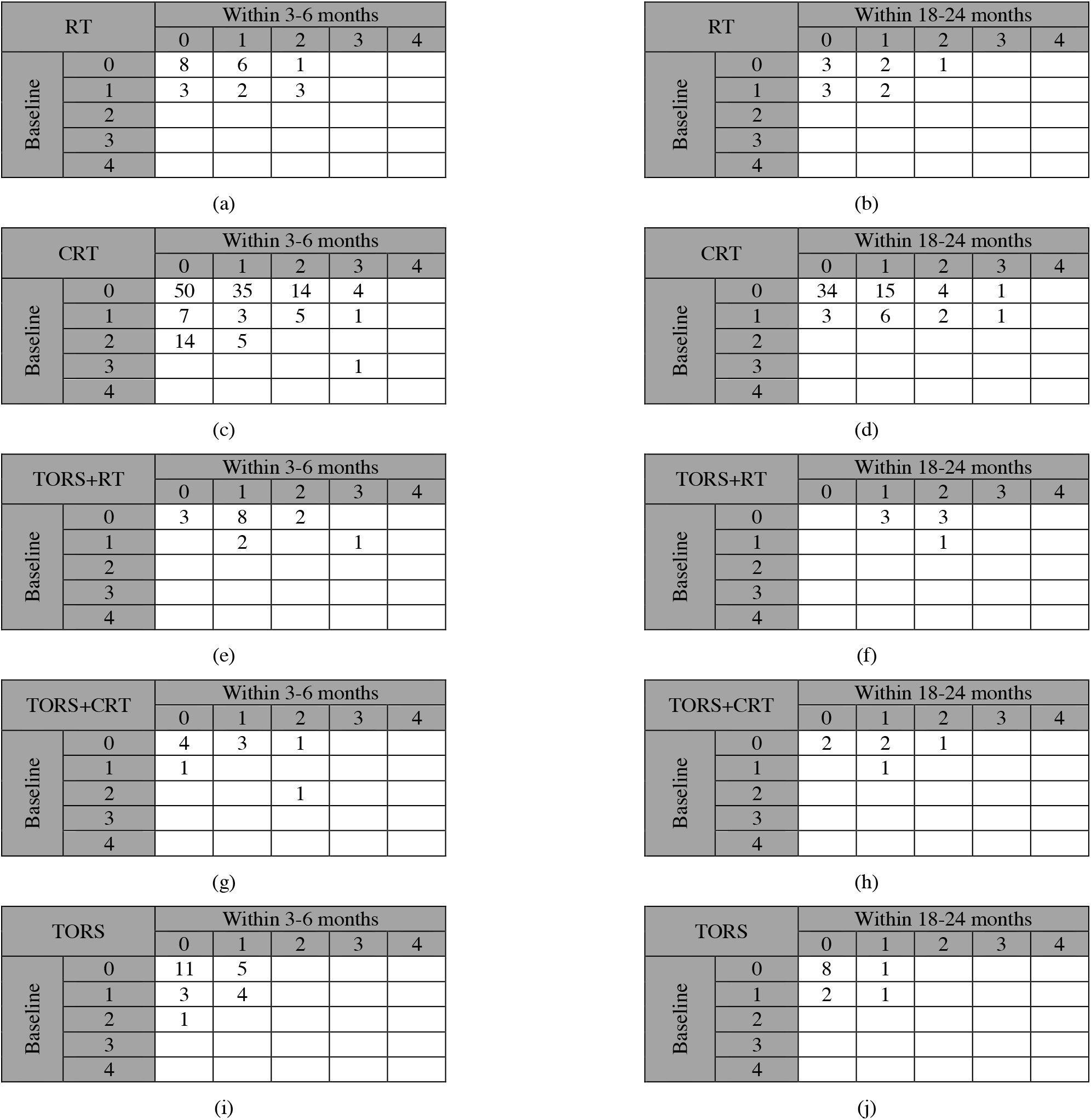
The evolution of DIGEST baseline grades within 3-6 moths and within 18-24 months post therapy.

| RT |  | Within 3-6 months |  |  |  |  |
| --- | --- | --- | --- | --- | --- | --- |
|  |  | 0 | 1 | 2 | 3 | 4 |
| Baseline | 0 | 8 | 6 | 1 |  |  |
|  | 1 | 3 | 2 | 3 |  |  |
|  | 2 |  |  |  |  |  |
|  | 3 |  |  |  |  |  |
|  | 4 |  |  |  |  |  |

(a)
| RT |  | Within 18-24 months |  |  |  |  |
| --- | --- | --- | --- | --- | --- | --- |
|  |  | 0 | 1 | 2 | 3 | 4 |
| Baseline | 0 | 3 | 2 | 1 |  |  |
|  | 1 | 3 | 2 |  |  |  |
|  | 2 |  |  |  |  |  |
|  | 3 |  |  |  |  |  |
|  | 4 |  |  |  |  |  |

(b)
| CRT |  | Within 3-6 months |  |  |  |  |
| --- | --- | --- | --- | --- | --- | --- |
|  |  | 0 | 1 | 2 | 3 | 4 |
| Baseline | 0 | 50 | 35 | 14 | 4 |  |
|  | 1 | 7 | 3 | 5 | 1 |  |
|  | 2 | 14 | 5 |  |  |  |
|  | 3 |  |  |  | 1 |  |
|  | 4 |  |  |  |  |  |

(c)
| CRT |  | Within 18-24 months |  |  |  |  |
| --- | --- | --- | --- | --- | --- | --- |
|  |  | 0 | 1 | 2 | 3 | 4 |
| Baseline | 0 | 34 | 15 | 4 | 1 |  |
|  | 1 | 3 | 6 | 2 | 1 |  |
|  | 2 |  |  |  |  |  |
|  | 3 |  |  |  |  |  |
|  | 4 |  |  |  |  |  |

(d)
| TORS+RT |  | Within 3-6 months |  |  |  |  |
| --- | --- | --- | --- | --- | --- | --- |
|  |  | 0 | 1 | 2 | 3 | 4 |
| Baseline | 0 | 3 | 8 | 2 |  |  |
|  | 1 |  | 2 |  | 1 |  |
|  | 2 |  |  |  |  |  |
|  | 3 |  |  |  |  |  |
|  | 4 |  |  |  |  |  |

(e)
| TORS+RT |  | Within 18-24 months |  |  |  |  |
| --- | --- | --- | --- | --- | --- | --- |
|  |  | 0 | 1 | 2 | 3 | 4 |
| Baseline | 0 |  | 3 | 3 |  |  |
|  | 1 |  |  | 1 |  |  |
|  | 2 |  |  |  |  |  |
|  | 3 |  |  |  |  |  |
|  | 4 |  |  |  |  |  |

(f)
| TORS+CRT |  | Within 3-6 months |  |  |  |  |
| --- | --- | --- | --- | --- | --- | --- |
|  |  | 0 | 1 | 2 | 3 | 4 |
| Baseline | 0 | 4 | 3 | 1 |  |  |
|  | 1 | 1 |  |  |  |  |
|  | 2 |  |  | 1 |  |  |
|  | 3 |  |  |  |  |  |
|  | 4 |  |  |  |  |  |

(g)
| TORS+CRT |  | Within 18-24 months |  |  |  |  |
| --- | --- | --- | --- | --- | --- | --- |
|  |  | 0 | 1 | 2 | 3 | 4 |
| Baseline | 0 | 2 | 2 | 1 |  |  |
|  | 1 |  | 1 |  |  |  |
|  | 2 |  |  |  |  |  |
|  | 3 |  |  |  |  |  |
|  | 4 |  |  |  |  |  |

(h)
| TORS |  | Within 3-6 months |  |  |  |  |
| --- | --- | --- | --- | --- | --- | --- |
|  |  | 0 | 1 | 2 | 3 | 4 |
| Baseline | 0 | 11 | 5 |  |  |  |
|  | 1 | 3 | 4 |  |  |  |
|  | 2 | 1 |  |  |  |  |
|  | 3 |  |  |  |  |  |
|  | 4 |  |  |  |  |  |

(i)
| TORS |  | Within 18-24 months |  |  |  |  |
| --- | --- | --- | --- | --- | --- | --- |
|  |  | 0 | 1 | 2 | 3 | 4 |
| Baseline | 0 | 8 | 1 |  |  |  |
|  | 1 | 2 | 1 |  |  |  |
|  | 2 |  |  |  |  |  |
|  | 3 |  |  |  |  |  |
|  | 4 |  |  |  |  |  |

